# Pre-diabetes in a real-world endocrine/diabetes clinic registry in Najaf, Iraq: a retrospective registry-based analysis

**DOI:** 10.64898/2026.02.05.26345696

**Authors:** Zeid Faiek Zwain

## Abstract

Prediabetes is a high-risk dysglycemic state. We used a real-world endocrine/diabetes clinic registry from Najaf, Iraq to characterize patients labeled as having pre-diabetes and to explore factors associated with follow-up engagement. We identified prediabetes visits using keyword-based case finding (English and Arabic terms including prediabetes/pre-diabetes, IFG, IGT, and impaired fasting glucose/tolerance) across semi-structured registry fields. Visit-level data were collapsed to patient-level records. Binary indicators of hypertension, dyslipidemia/statin use, obesity/weight management, smoking, and common glucose-lowering therapies were derived from registry text using keyword/brand-name matching. The primary outcome was follow-up engagement defined as ≥2 recorded visits. The prediabetes subset comprised 242 unique patients and 302 visits. Median age was 45 years (IQR 35–55); 47 patients (19.4%) had ≥2 visits. Median follow-up duration was 0 days (maximum 321). Obesity/weight-management indicators were frequent (71.1%), as were hypertension (43.4%) and dyslipidemia/statin indicators (46.3%).

In multivariable logistic regression, no evaluated predictor reached conventional statistical significance for follow-up engagement. Registry enhancements to capture laboratory confirmation and standardized follow-up fields may improve the ability to evaluate diabetes prevention pathways.

## Introduction

Prediabetes is defined by impaired fasting glucose, impaired glucose tolerance, or elevated HbA1c below the diagnostic threshold for diabetes. It is associated with increased risk of progression to type 2 diabetes and cardiometabolic disease. Trials such as the Diabetes Prevention Program demonstrated that intensive lifestyle intervention and metformin reduce progression to diabetes in high-risk individuals. Real-world clinic registries can support surveillance and evaluation of prevention pathways, but their utility depends on standardized diagnostic capture and longitudinal follow-up.

We aimed to (1) characterize patients labeled as having prediabetes in a real-world clinic registry in Najaf, Iraq, and (2) explore factors associated with follow-up engagement (≥2 visits).

## Methods

### Study design and setting

Retrospective observational registry-based study using routine-care data from Zeid Zwain Clinic (Najaf, Iraq).

### Data source

The clinic registry contains visit-level records including demographics, visit date, and semi-structured text fields (summary, past history, drugs, investigations).

### Prediabetes case identification

Prediabetes visits were identified using keyword-based case finding across text fields with English and Arabic terms (including prediabetes/pre-diabetes, IFG, IGT, impaired fasting glucose, impaired glucose tolerance, and Arabic equivalents). This approach captures registry labeling and may not always correspond to laboratory-confirmed definitions.

### Derivation of indicators

We derived binary indicators from the semi-structured text using keyword and brand-name matching for: hypertension, dyslipidemia/statin use, obesity/weight-management, smoking, metformin, GLP-1 receptor agonists, and SGLT2 inhibitors.

### Outcome

The primary outcome was follow-up engagement, defined as having ≥2 recorded visits in the registry among identified prediabetes patients.

### Statistical analysis

We summarized categorical variables as counts and percentages and continuous variables as median (IQR). We fitted multivariable logistic regression models to estimate adjusted odds ratios for follow-up engagement. Analyses were performed in Python.

### Ethics and data availability

Ethics approval obtained by local authority before starting study. Data availability: de-identified data and documentation are available at Zenodo DOI 10.5281/zenodo.18239322.

## Results

Prediabetes case-finding identified 242 unique patients and 302 visits. Median age was 45 years (IQR 35–55). Follow-up engagement (≥2 visits) occurred in 47 patients (19.4%), with median follow-up 0 days (maximum 321). Figure 1 shows the distribution of visit counts per patient.

**Figure 1.**
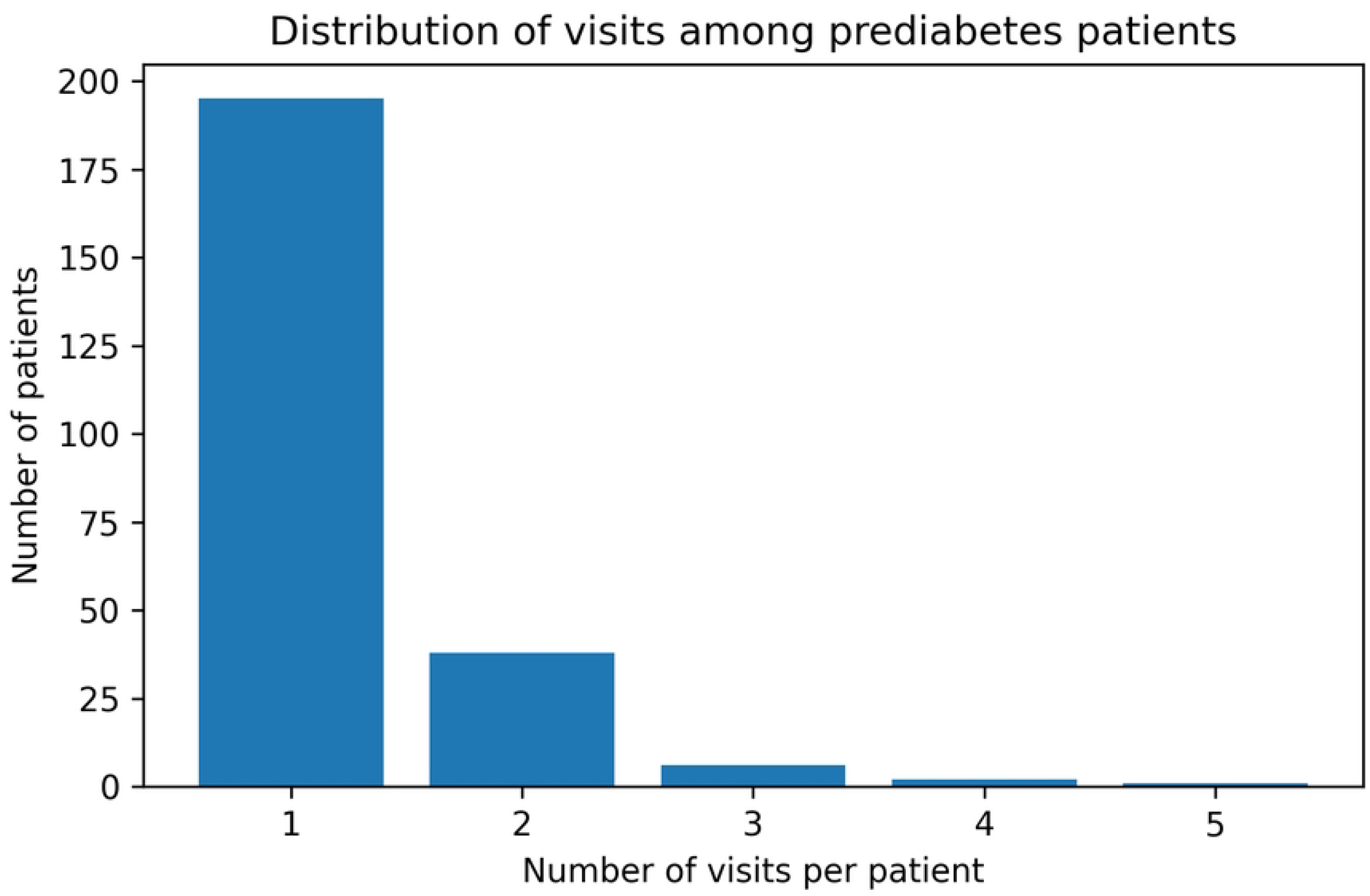
Distribution of visits among prediabetes patients. Bar chart showing the number of patients by total number of recorded visits in the clinic registry.

Comorbidity and therapy indicators (patient-level prevalence):

Figure 2 shows the prevalence of selected comorbidity and therapy indicators in the prediabetes cohort.

**Figure 2.**
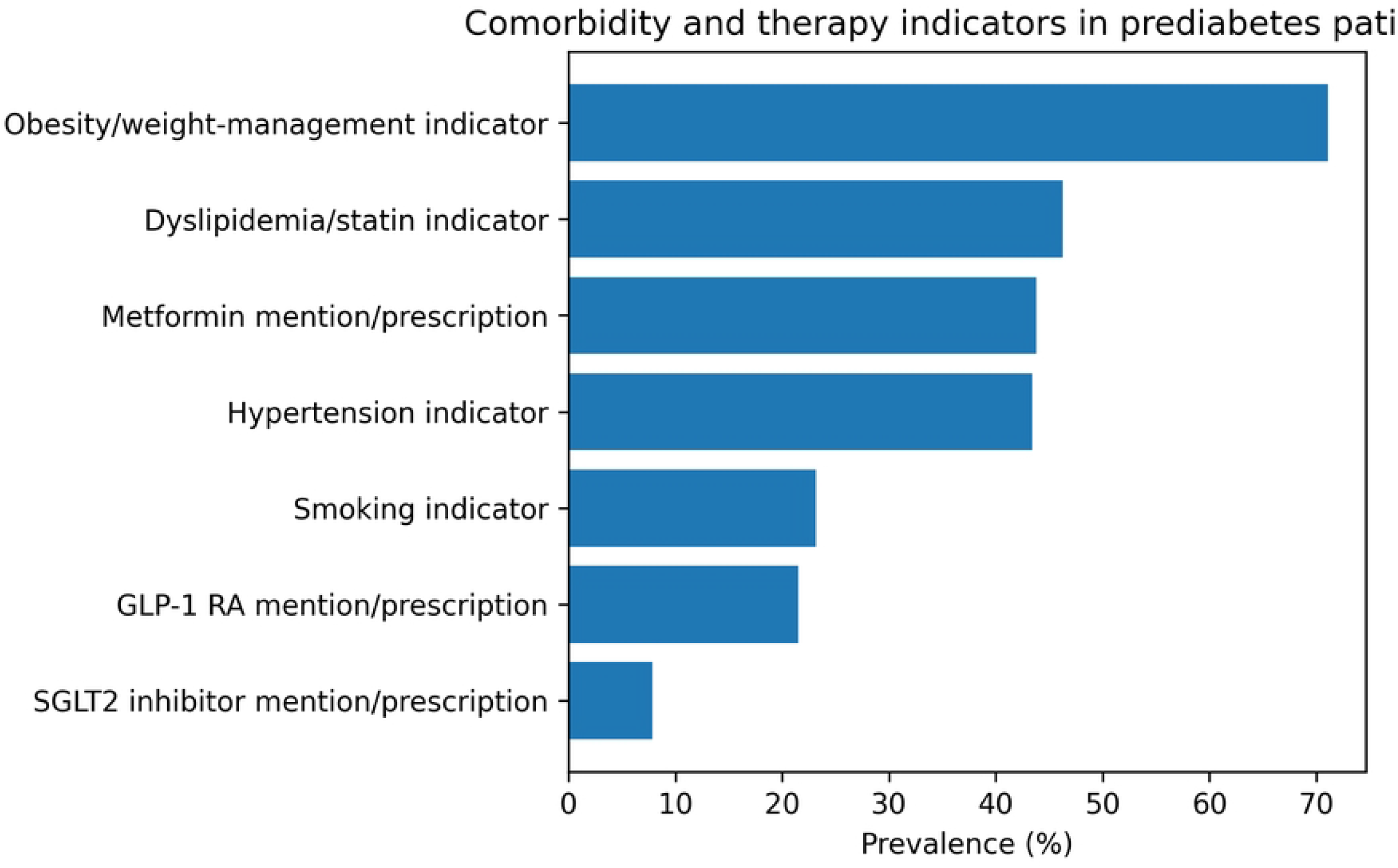
Comorbidity and therapy indicators in prediabetes patients. Horizontal bar chart showing patient-level prevalence (%) of text-derived indicators: obesity/weight-management, dyslipidemia/statin, hypertension, metformin, smoking, GLP-1 receptor agonist, and SGLT2 inhibitor.

- Obesity/weight-management indicator: 71.1%
- Dyslipidemia/statin indicator: 46.3%
- Metformin mention/prescription: 43.8%
- Hypertension indicator: 43.4%
- Smoking indicator: 23.1%
- GLP-1 RA mention/prescription: 21.5%
- SGLT2 inhibitor mention/prescription: 7.9%

## Discussion

In this single-clinic real-world registry, prediabetes labeling identified a modest subset of patients, with limited longitudinal follow-up for most individuals. Indicators consistent with obesity/weight-management and cardiometabolic comorbidities were frequent. Follow-up engagement was observed in about one in five patients; in exploratory adjusted analyses, we did not identify statistically significant predictors of follow-up.

These findings support practical improvements to registry structure, including systematic capture of diagnostic laboratories (HbA1c, fasting glucose, OGTT where relevant), standardized coding, and structured follow-up variables (planned review date, lifestyle intervention referral, and prevention pharmacotherapy when indicated).

## Limitations

Prediabetes ascertainment relied on keyword-based registry labeling rather than uniform laboratory confirmation. Indicators were derived from semi-structured text and may be misclassified. This is a single-clinic registry and may not be generalizable. Sex coding in the identified subset should be verified in the underlying registry and improved in future iterations.

## Conclusions

A real-world clinic registry can identify prediabetes populations, but standardized biochemical capture and longitudinal follow-up fields are needed to support robust evaluation of diabetes prevention pathways. This will bring local authority attention to the increasing risk of prediabetes in the community so as to put prevention criteria.

## Data Availability

All de-identified data underlying the findings of this study, together with the data dictionary and documentation, are publicly available in the Zenodo repository at DOI: 10.5281/zenodo.18239322

https://doi.org/10.5281/zenodo.18239322

## Acknowledgments

None

## References

1. Knowler WC, Barrett-Connor E, Fowler SE, et al. Reduction in the incidence of type 2 diabetes with lifestyle intervention or metformin. N Engl J Med. 2002;346:393–403.

2. American Diabetes Association. Standards of Care in Diabetes (Diagnosis and Classification of Diabetes). [Year].

3. von Elm E, Altman DG, Egger M, et al. The STROBE statement. Lancet. 2007;370:1453–1457.

